# Characterizing relevant microRNA editing sites in Parkinson’s disease

**DOI:** 10.1101/2020.04.06.20054791

**Authors:** Chenyu Lu, Shuchao Ren, Zhigang Zhao, Xingwang Wu, Angbaji Suo, Nan Zhou, Jun Yang, Shuai Wu, Tianqing Li, Chao Peng, Yun Zheng

## Abstract

MicroRNAs (miRNAs) are extensively edited in human brains. However, the functional relevance of miRNA editome is largely unknown in Parkinson’s disease (PD). By analyzed small RNA sequencing profiles of brain tissues of 43 PD patients and 88 normal controls, we totally identified 421 miRNA editing sites with significantly different editing levels in prefrontal cortices of PD patients (PD-PC). A-to-I edited miR-497-5p has significantly higher expression levels in PD-PC compared to normal controls and directly represses OPA1 and VAPB, which potentially contributes to the progressive neurodegeneration of PD patients. These results provide new insights into mechanistic understanding, novel diagnostic and therapeutic clues of PD.

## Introduction

MicroRNAs (miRNAs) are small non-coding RNAs with about 22 nucleotides that normally repress their target mRNAs at post-transcriptional level [1]. Some miRNAs are edited, such as Adenosine-to-Inosine (A-to-I) editing [2–7] performed by adenosine deaminase (ADAR) enzymes and C-to-U editing performed by apolipoprotein B mRNA editing catalytic polypeptide-like (APOBEC) enzymes [8], during their biogenesis processes. Altered editing of miRNAs lead to human diseases, such as cancer [9–11]. Parkinson’s disease is a neurodegenetative disorder affecting 2-3% of people over 65 years of age [12]. However the functional relevance of miRNA editing in PD is largely unknown, although A-to-I editing is prevalent in brain [13–16] and the editing level is gradually increasing in the developmental procedure [5,16].

To comprehensively characterize miRNA editing sites in brain tissues of PD, we analyzing 43 and 88 small RNA sequencing profiles of PD and control brain tissues, respectively. We identified 421 miRNA editing sites that have significantly different editing levels in prefrontal cortices of PD patients (PD-PC) compared with control group. The editing levels of 6 A-to-I and 3 C-to-U editing sites are significantly correlated with the ages of normal controls which is disrupted in PD patients. One A-to-I editing site in miR-497-5p has significantly higher editing levels compared to normal controls in PD-PC samples and edited miR-497-5p directly represses OPA1 and VAPB, which potentially contributes to the progressive neurodegeneration of PD patients. These results demonstrate that miRNA editing is severely disturbed and relevant in PD and offer novel insight into the etiology of PD. The editing of miRNAs might be used to develop novel diagnostic biomarkers and/or therapeutic targets for PD.

## Results

### Summary of data sets used

To comprehensively identify miRNA mutation and editing (M/E) sites in PD, we collected 131 sRNA-seq profiles of postmortem PD patients and normal people from NCBI SRA Database (Supplementary Table S1.1). These profiles include 29 prefrontal cortex samples of PD patients (PD-PC), 14 amygdalae samples of PD patients (PD-Am), 36 prefrontal cortex samples of normal controls (PC), 14 amygdalae samples of normal controls (Am), 6 frontal cortex samples of normal controls (FC), 6 corpus callosum samples of normal controls (CC), 2 inferior parietal lobe samples of normal controls (IPL), 2 temporal neocortex gray matter samples of normal controls (NG), 3 astrocyte cell lines of normal controls (As) and 19 unknown brain regions of normal controls (Unknown). The PD-PC and PC samples were from Brodmann Area 9 (BA9). To understand the potential function of edited miRNAs, we examined the deregulated genes and proteins from the BA9 and several other brain regions of PD patients (Table S1.2).

### An overview of identified editing sites in miRNAs

We used the MiRME pipeline [17] to analyze the 131 sRNA-seq profiles selected with the default settings. Totally, we found 2456 significant M/E sites (as shown in Figure 1 and Table S2.1), by the criteria of at least 10 reads and multiple test corrected *P* -values smaller than 0.05. The largest category of these M/E sites are 3’-A, accounting for 37.3% (916 sites), then followed by 3’-U (28.6%), 3’-Other (15.5%) and 5’-editing (5.4%) (Fig. 1a). A-to-I and C-to-U editing sites account for 1.9% (46 sites) and 1.1% (26 sites), respectively (Fig. 1a). We found 60 SNPs after comparing the identified M/E sites to SNPs in dbSNP (Fig. 1a). Other types of editing sites consist of 6.7% (164 sites) (Fig. 1a).

**Fig. 1.**
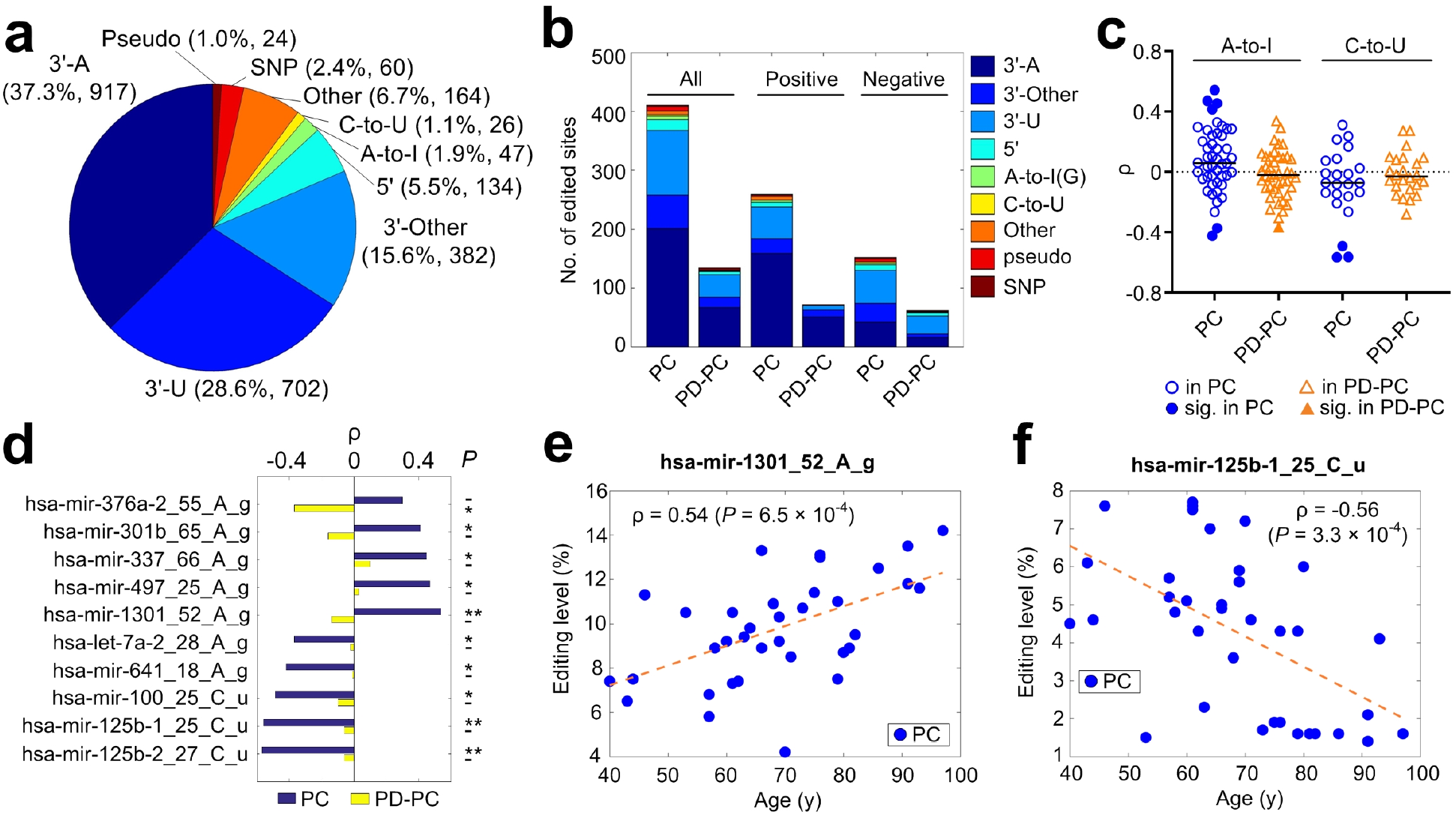
A summary of the identified miRNA mutation and editing sites. (a) The categories of significant M/E sites in miRNAs. (b) The numbers of different types of editing sites that have significant Spearman correlation (*ρ*) between editing levels and the ages of individuals in the PC and PD-PC group. (c) The distributions of *ρ* for A-to-I and C-to-U sites in the PC and PD-PC samples, respectively. (d) Ten selected editing sites with significant *ρ* values in either PC or PD-PC samples. *: *P* < 0.05; **: corrected *P* < 0.05. (e) to (f) The plots of ages of PC samples against the editing levels of two selected editing sites. See also Extended Data Fig. 1.

The A-to-I, C-to-U and Other sites were further classified based on their types of variations (Extended Data: Fig. 1a). Similar to previous work [17], there could be several 3’-editing sites in one pre-miRNA, but most pre-miRNAs only have 1 or 2 central sites in our selected samples (ED: Fig. 1b). However, some miRNAs may have a few editing events at 5’ end in our previous study [17].

### Age-related editing sites in PD and normal controls

Because previous studies reported that A-to-I editing level is increasing during the development process [5,16], we examined the Spearman correlation (*ρ*) between editing levels of the 2456 editing sites and the ages (at death) of 36 PC and 29 PD-PC samples, respectively. There are 411 sites with significant *ρ* in PC, but only 134 sites with significant *ρ* in PD-PC samples (Fig. 1b and Table S2.2-2.3). When these sites were separated as ones with positive and negative *ρ* values, the numbers of sites with significant *ρ* values in PD-PC samples are also much smaller than those in PC samples (Fig. 1b and ED: Fig. 1c). The numbers of types of editing sites are decreased in PD-PC samples too. For example, there are no A-to-I sites with positive *ρ* values in the PD-PC samples (ED: Fig. 1c). Furthermore, the Kullback-Leibler divergences of the distributions of different types of editing sites in PC and PD-PC samples are 0.6, 1.9 and 1.7 for all, sites with positive *ρ* values and negative *ρ* values, respectively (ED: Fig. 1c). These results suggest that the gradually increasing or decreasing editing levels of miRNAs during the aging procedure are severely disturbed in PD.

The *ρ* values of 47 A-to-I and 26 C-to-U editing sites are carefully examined in Fig. 1c. In PC samples, there are 4 and 2 A-to-I sites with significant positive and negative *ρ* values, respectively (Fig. 1c-1e). In comparison, only one A-to-I site has significant negative *ρ* in PD-PC (Fig. 1c-1d). The median value of *ρ* in PC is slightly larger than 0, which is consistent with ever-increasing A-to-I editing level along age noticed previously [5,16]. But the median *ρ* value in PD-PC samples is smaller than 0, further suggesting the disrupted A-to-I editing of miRNAs in PD. In contrast, the C-to-U editing sites have a weak negative median *ρ* value in PC, suggesting C-to-U editing sites of miRNAs generally have decreasing editing levels when people are aging. Three C-to-U editing sites (see Fig. 1c, 1d and 1f) have significant *ρ* value in PC.

### A-to-I editing sites

We totally found 47 significant A-to-I editing sites from the selected samples (ED:Figure 2a and Table S2.4). Among them, 14 sites are newly identified when compared with reported sites (ED: Fig. 2a). At least 19 of these 47 sites are conserved in other mammals [3,18] (ED: Fig. 2a). Similar to previous results [3,4,17,18], the 5’ and 3’ side of these A-to-I editing sites preferred to be U and G, respectively (ED: Fig. 2b). For examples, two A-to-I editing sites were shown in ED: Fig. 2c to 2f.

**Fig. 2.**
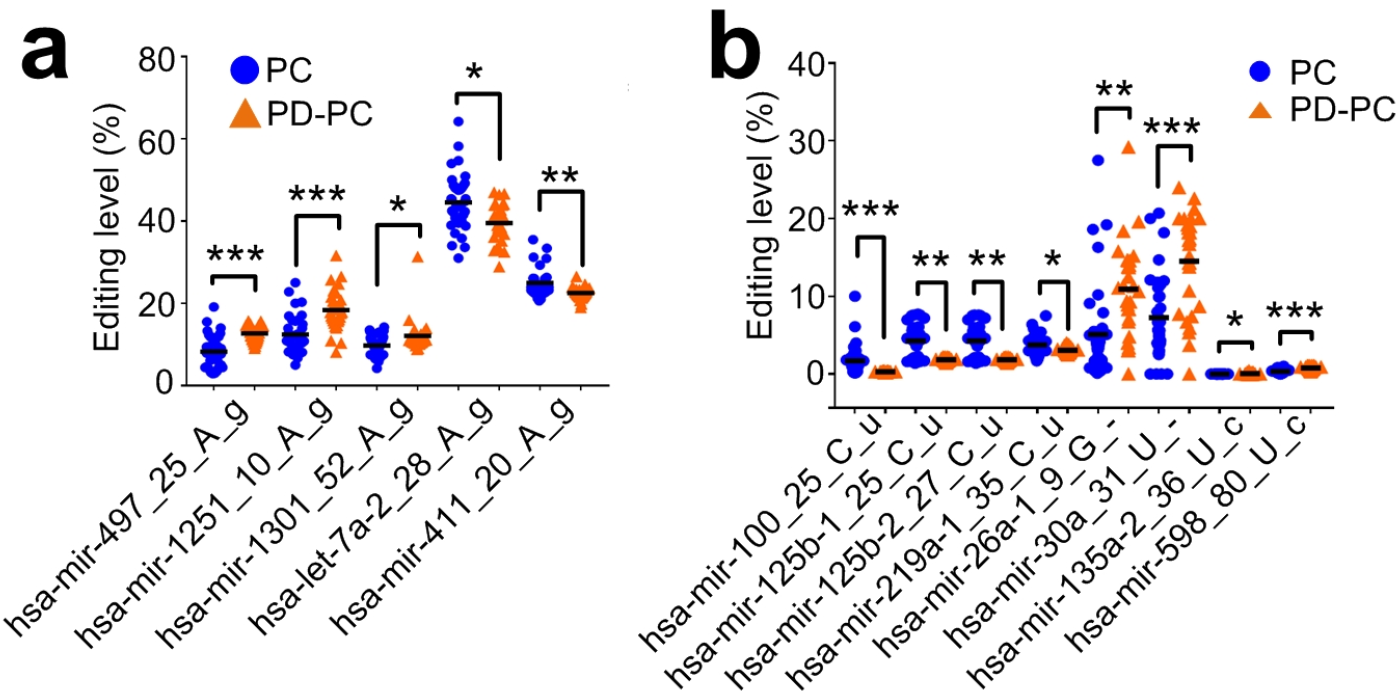
M/E sites that have significantly different editing levels in PD-PC. (a) Five A-to-I editing sites with significantly different editing levels in PD-PC. (b) Eight editing sites with significantly different editing levels in PD-PC. *: corrected *P* < 0.05, **: corrected *P* < 0.01, ***: corrected *P* < 0.001, Mann-Whitney *U* -test. See also Extended Data Fig. 6a-6b.

### C-to-U editing sites

We carefully explored the 26 C-to-U editing sites (ED: Fig. 3a and Table S2.5). Five of these 26 C-to-U editing sites are conserved in primates [18] (see ED: Fig. 3a). The neighboring nucleotides of these C-to-U sites prefer to be C on both 5’ and 3’ sides (ED: Fig. 3b), consistent with the CCC motif of APOBEC3G [19]. Three examples of C-to-U editing sites are shown in ED: Fig. 3c-3h. There are multiple C-to-U editing sites on two pre-miRNAs, i.e., hsa-mir-219a-2 and hsa-mir-1249 (ED: Fig. 4 and Table S2.5). Interestingly, seven continuous cytosines on hsa-mir-1249 were found to be C-to-U edited (ED: Fig. 3a and Fig. 4).

**Fig. 3.**
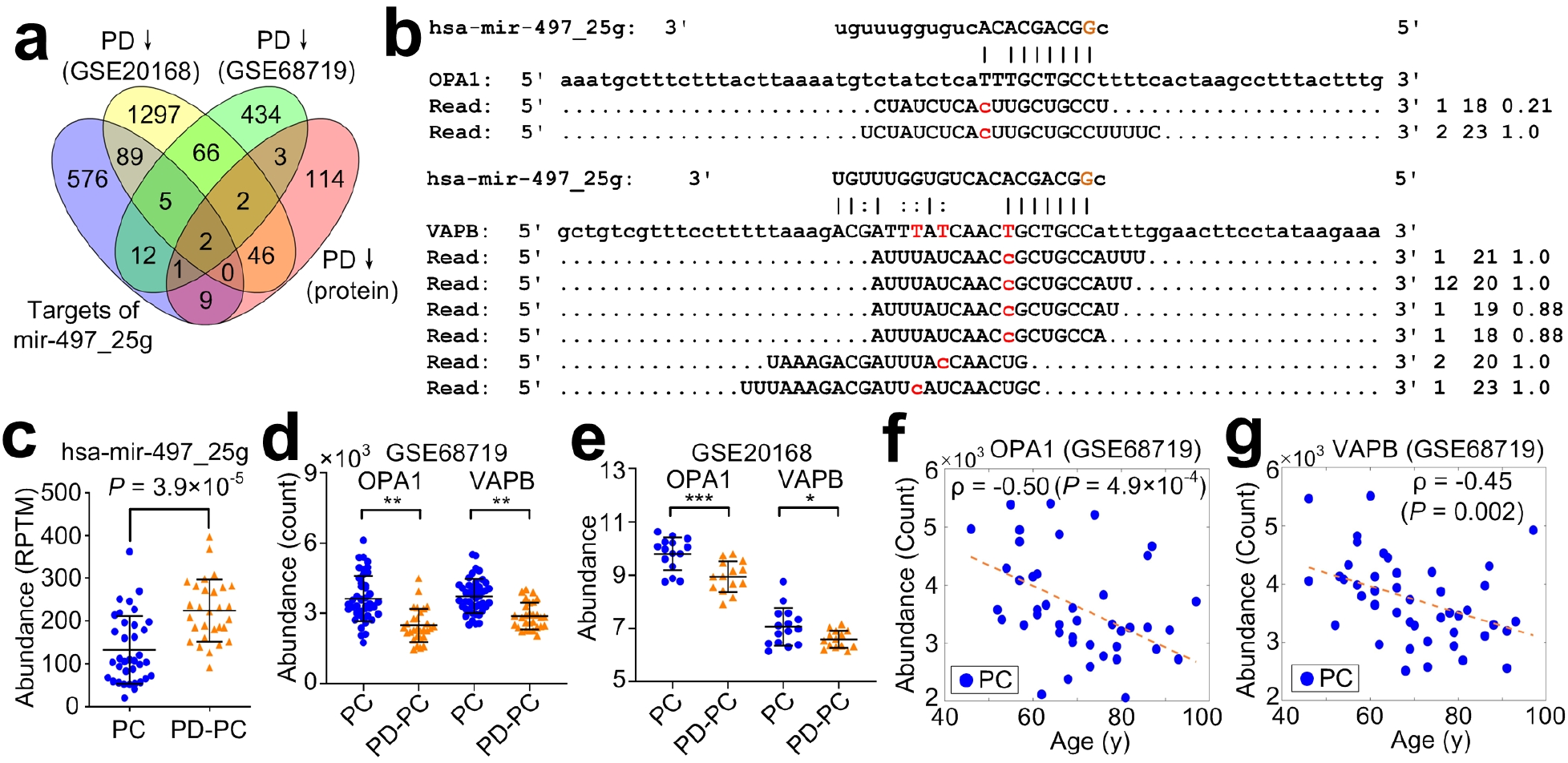
The target analysis of A-to-I edited hsa-miR-497-5p. (a) Integrated analysis of targets of edited hsa-miR-497-5p (hsa-mir-497_25g) and the downregulated genes and proteins in PD-PC (BA9). (b) The complementary sites of hsa-mir-497_25g on OPA1 and VAPB, and the PAR-CLIP sequencing reads from these sites. (c) Comparison of abundances of hsa-mir-497_25g in PC and PD-PC. (d) - (e) Comparison of abundances of OPA1 and VAPB in PC and PD-PC (BA9). *: *P* < 0.05; **: corrected *P* < 0.05; ***: corrected *P* < 0.001, DESeq2 and limma package, respectively. (f) - (g) The plots of the ages of PC samples against the abundances of OPA1 and VAPB. See also Extended Data Fig. 7 and 8.

**Fig. 4.**
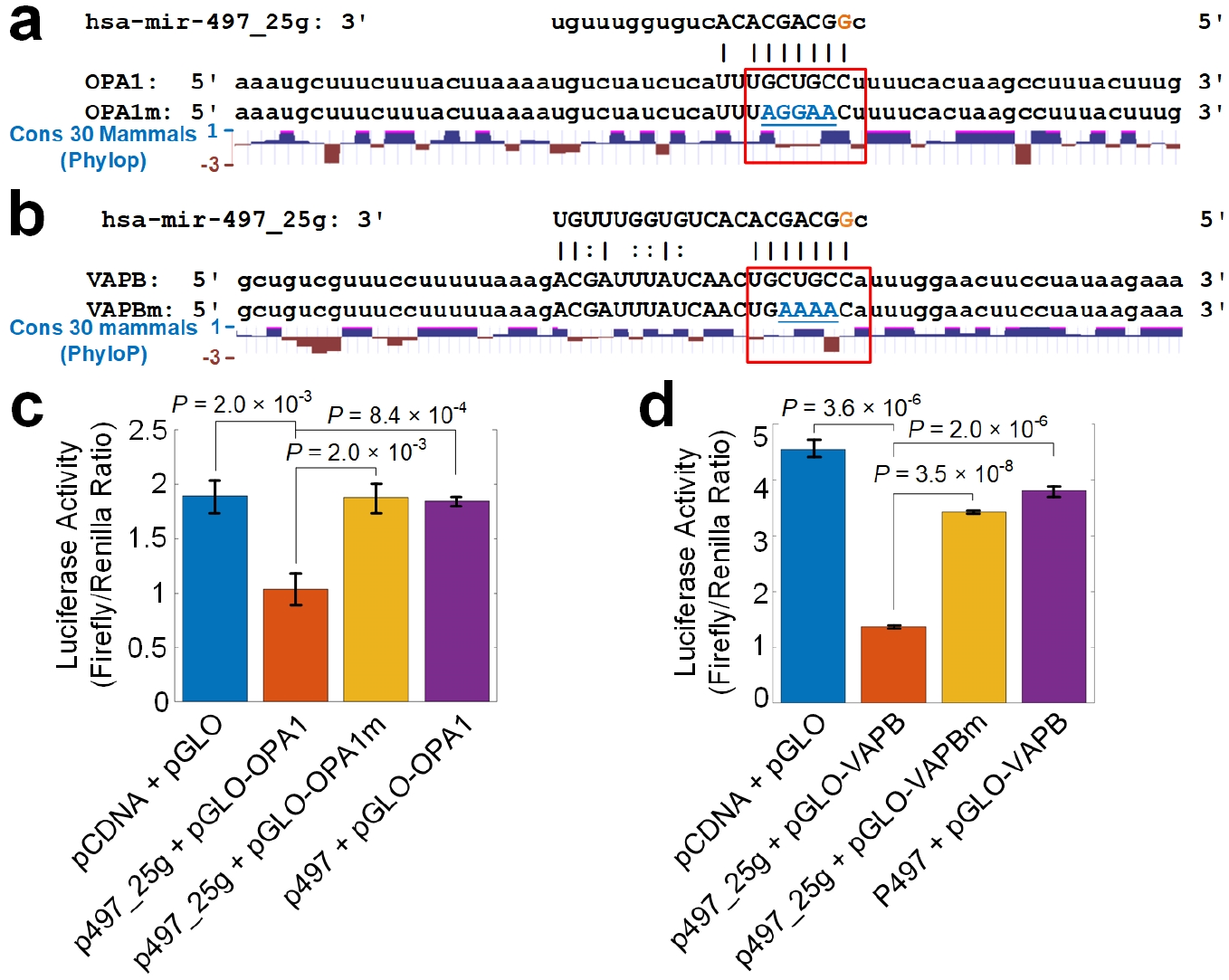
Validating selected targets of A-to-I edited miR-497-5p. (a) - (b) The segment of 3’ UTR of OPA1 and VAPB, mutated segment of 3’ UTR of OPA1 and VAPB, segment of 3’ UTR of mouse Opa1 and Vapb, and conservation scores for 30 mammals in these region calculated with PhyloP, resepctively. (c) - (d) The luciferase activities when co-transfecting a pGLO plasmid of 3’ UTRs in Part (a) - (b) and a pCDNA plasmid containing original pre-hsa-mir-497 (p497) or pre-hsa-mir-497_25g (p497 25g), respectively. The values shown are mean values *±* SDs. *P* -values were based on two-tailed *t*-tests. See also Extended Data Fig. 9.

### Identified SNPs in miRNAs

By comparing the M/E sites to SNPs reported in dbSNP and examining their editing levels, 60 M/E sites were regarded as SNPs (as shown in ED: Fig. 5a and Table S2.6). Three of the 60 SNP sites were shown in ED: Fig. 5b to 5g. The editing levels of these three sites are 100% in the selected samples (ED: Fig. 5e to 5g).

### Relevant miRNA editing sites in PD

To identify editing sites that are relevant in PD, we compared the editing levels of 2456 significant editing sites in PD-PC and PC; PD-Am and Am samples, respectively. We obtained 421 M/E sites that have significantly different editing levels in PD-PC samples compared to PC samples (ED: Fig. 6a and Table S3). Most of 421 sites (65.8% or 277 sites) have increased editing levels in PD-PC samples, and 34.2% of these sites have decreased editing levels in PD-PC samples (ED: Fig. 6b). Three A-to-I editing sites (hsa-mir-497_25_A_g, hsa-mir-1251_10_A_g, and hsa-mir-1301_52_A_g) have increased editing levels in PD-PC samples, and two A-to-I editing sites (hsa-let-7a-2_28_A_g and hsa-mir-411_20_A_g) show decreased editing levels in PD-PC samples (Fig. 2a). Four C-to-U and four Other editing sites have decreased editing levels in PD-PC samples (Fig. 2b). There are no M/E sites with significantly different editing levels in the comparison between PD-Am and Am samples.

### Target analysis of A-to-I editing sites in seeds

We selected 4 M/E sites that locate in the seed regions of mature miRNAs (Table S3) and predicted their targets with the MiCPAR pipeline [20], as listed in Table S4.1. After comparing the results to those of original miRNAs (in Table S4.2), we found that these editing events severely changed the target sets of these miRNAs (ED: Fig. 6c), as well as the enriched GO terms and KEGG pathways of these targets (ED: Fig. 6c and Table S5).

Since A-to-I edited hsa-miR-497-5p (hsa-mir-497_25g) has much higher expression level than the other three A-to-I edited miRNAs (ED: Fig. 6d), we next carefully examined the targets of hsa-mir-497_25g. After comparing the predicted targets of edited miRNAs with the deregulated genes and proteins in prefrontal cortex (BA9) samples (Fig. 3a, see details in Methods), we found that hsa-mir-497_25g targets OPA1 mitochondrial dynamin like GT-Pase (OPA1) and VAMP associated protein B and C (VAPB) (Fig. 3b). Presumably due to the upregulation of hsa-mir-497_25g (Fig. 3b) in PD-PC, OPA1 and VAPB are significantly downregulated in PD-PC (Fig. 3d-3e). Furthermore, the protein expression level of OPA1 and VAPB is also significantly downregulated in PD-PC samples (corrected *P* < 0.05, as in [21]).

Furthermore, OPA1 is significantly downregulated in substantia nigra (SN) (ED: Fig. 7a-7b), is mildly decreasing in putamen of PD patients (ED: Fig. 7c). But in subventricular zone (SZ) and cingulate gyrus, the expression of OPA1 does not change significantly (ED: Fig. 7d-7e). In SN, VAPB has a weak decreasing trend in PD (ED: Fig. 7a), but has no change in another cohort (ED: Fig. 7b), and cingulate gyrus, SZ, putamen (ED: Fig. 7c to 7e, respectively).

Importantly, consistent with the significant positive correlation between editing level of hsa-mir-497_25_A_g and ages of individuals in PC (Fig. 2f), the expression levels of OPA1 and VAPB are significantly negatively correlated with the ages of individuals in PC (Fig. 3f, 3g and ED: Fig. 7f/8f) and potentially in SN as well (*ρ* = *−*0.36 and *−*0.47, respectively, see ED: Fig. 7g and 8g, respectively). The Spearman correlation *ρ* between expression of OPA1 and VAPB and ages in putamen are weak and insignificant in putamen (ED: Fig. 7h and 8h, respectively), and are positive in SZ (*ρ* = 0.29 and 0.60 in ED: Fig. 7i and 8i, respectively). In PD, these correlations were generally weakened (ED: Fig. 7j-7l/7n and 8j-8l/8n for OPA1 and VAPB, respectively), except in putamen where the negative correlations are slightly enhanced (ED: Fig. 7m and 8m for OPA1 and VAPB, respectively).

### hsa-mir-497_25g directly represses OPA1 and VAPB

We first examined the conservation of the complementary site of hsa-mir-497_25g on VAPB. The 7mer opposite to the seed of edited miR-497-5p is only partially conserved in mammals (Fig. 4a). Especially, the 3rd nucleotide from 5’ end (as pointed by a red arrow in Fig. 4a) has a conservation score of -1.47, indicating that the site is fast-evolving in primates. Even within primates, 7 species did not have conserved sequences complementary to the seed of hsa-mir-497_25g (Fig. 4b).

To verify that hsa-mir-497_25g directly represses OPA1 and VAPB, we performed luciferase assays in 293T cells for hsa-mir-497_25g, original hsa-mir-497, OPA1/VAPB, mutated OPA1/VAPB (see Fig. 4a and 4b, respectively). As in Fig. 4c and 4d, hsa-mir-497_25g induced significant decreases of luciferase activities for OPA1 and VAPB, indicating that hsa-mir-497_25g directly represses OPA1 and VAPB, respectively. In comparison, when being co-transfected with mutated OPA1 and mutated VAPB, hsa-mir-497_25g could not repress mutated OPA1 and mutated VAPB (Fig. 4c and 4d, respectively). The original miR-497-5p could not repress OPA1 and VAPB too (Fig. 4c and 4d, respectively). The complementary sites of hsa-mir-497_25g on OPA1 and VAPB have non-conserved seed regions (see red rectangles in Fig. 4a-4b). Actually, these two hsa-mir-497_25g complementary sites are only conserved in some primates including monkey, but non-conserved in rodents (ED: Fig. 9a-9d). As expected, A-to-I edited miR-497-5p could repress monkey OPA1 and VAPB, but could not repress mouse Opa1 and Vapb (ED: Fig. 9e-9h).

To summarize, hsa-mir-497_25g directly represses OPA1 and VAPB in human, and putatively in some primates, but not in mouse and those primates with non-conserved complementary sites of hsa-mir-497_25g, which might contributes to the advanced functions and fast evolving of prefrontal cortices in human.

## Discussion

*OPA1* localizes to the inner mitochondrial membrane and plays roles in regulating mitochondrial stability and energy output. Mutations in OPA1 have been associated with optic atrophy type 1 [22,23]. Recently, the association between OPA1 and parkinsonism is also being noticed [24,25]. One recent study found that 3 members of a family had autosomal dominant optic atrophy caused by OPA1 mutations and 2 of them developed nonsyndromic PD [26]. OPA1 is downregulated in PD-PC (BA9) (Fig. 2d-2e) and in substantia nigra of PD patients (ED: Fig. 7b-7c). PD patients carrying the G2019S mutation showed decreased levels of mature OPA1 [27]. OPA1 haploinsufficinecy leads to progressive loss of iPSC-derived dopaminergic neurons [28]. Autosomal dominant LRRK2 mutations are associated with both familial and sporadic PD [27,29]. LRRK2 directly interacts with OPA1, presumably to regulate membrane dynamics important for mediating endocytosis and mitochondrial function [27].

Neuronal synaptic dysfunction is one of the key features of Parkinsons disease [30,31]. VAPB protein is a membrane protein found in plasma and intracellular vesicle membranes, and involved in vesicle trafficking. VAPB is downregulated in BA9 of PD-PC (Fig. 3d-3e). Down-regulation of VAPB and its interacting protein PTPIP51 reduces dentritic spine numbers and synaptic activity [32]. A missense mutation (P56S) in VAPB was reported to associated with motorneuron degeneration in affected amyotrophic lateral sclerosis (ALS) patients [33]. Transgenic mice that expressed human P56S *VAPB* developed progressive hyperactivities and other motor abnormalities, and showed progressive loss of corticospinal motor neurons [34]. In contrast, overexpression of VAPB in mouse ALS model slows the motor impairment and promotes the survival of spinal motor neurons [35].

α-synuclein is a PD-related protein and localizes to the nerve terminal [36] and directly binds to VAPB [36] and VAMP2 [37]. Disruption of the interaction between VAPB and PTPIP5 has been suggested to mediated the neurotoxic effect of α-synuclein. The downregulation of VAPB in PD patients might exaggerate the toxic effect of α-synuclein.

In summary, our results suggest that the editing level of hsa-mir-497_25_A_g increases in PD-PC samples and the A-to-I edited miR-497-5p dominantly represses OPA1 and VAPB in PD-PC samples, which leads to significant downregulation of OPA1 and VAPB in PD-PC. Consequently, the reduced OPA1 and VAPB contributes to the progressive neurodegeneration in PD. These results suggest that novel therapies of PD might be designed by either over-expressing OPA1/VAPB or repressing editing level of hsa-mir-497_25_A_g. Furthermore, the significant correlation between ages of normal people and editing level of hsa-mir-497_25_A_g, as well as expression levels of OPA1 and VAPB, and the significantly increased editing level of hsa-mir-497_25_A_g in PD might be used to develop novel diagnostic methods and therapies of PD.

## Methods

### The small RNA sequencing profiles used

As summarized in Supplementary Table S1.1, we collected 131 sRNA-seq profiles of post-mortem brain samples of PD patients and normal controls, including 29 prefrontal cortex samples of PD patients, 14 amygdalae samples of PD patients, 36 prefrontal cortex samples of normal controls, 15 amygdalae samples of normal controls, 6 frontal cortex samples of normal controls, 6 corpus callosum samples of normal controls, 3 astrocyte cell line samples of normal controls, 4 inferior parietal lobe samples of normal controls, 2 temporal neocortex gray matter samples of normal controls, and 17 unknown region samples of normal controls.

### The gene and protein expression profiles used

To understand the potential function of edited miRNAs, we examined the deregulated genes from brain samples of PD patients and normal controls. As summarized in Table S1.2, we chose 107 and 122 gene expression profiles of PD patients and normal controls, respectively, from 7 cohorts, including 2 cohorts for prefrontal cortex (Brodmann Area 9, briefly BA9) (GSE20168 and GSE68719), 2 cohorts for substantia nigra (GSE7621 and GSE20292), 1 cohort for putamen (GSE20291), 1 cohort for cingulate gyrus (GSE110716), and 1 cohort for subventricular zone (GSE130752).

We also checked 283 deregulated proteins in prefrontal contex (BA9) samples of PD by comparing the MS3 proteomics profiles of 12 PD patients to those of 12 normal people reported previously [21].

### Genome and annotation of miRNAs used

The unmasked genomic sequence of human (hg38, GRCh38) were downloaded from UCSC Genome Browser [38]. The bowtie-build program in the Bowtie package [39] was used to generate index files of human genome. The pre-miRNA sequences and genomic positions of miRNAs in gff3 format were downloaded from the miRBase (release 21) [40].

### Analysis of small RNA sequencing profiles

The selected sRNA-seq profiles were analyzed using the MiRME pipeline [17] with the default settings. The following criteria were used to define M/E sites with significant editing levels: (i) the relative level of editing is at least 5%; (ii) at least 10 reads support the editing event; (iii) the score threshold of sequencing reads is 30; and (iv) a multiple-test corrected *P* -value (using the Benjamini and Hochberg method [41]) of smaller than 0.05. Then, the obtained results of different samples were combined by a separate program in the MiRME package (see details in [17]). The identified M/E sites were compared to reported editing sites in miRNAs in the DARNED database [42], the RADAR database [43] and literature [3,44,4,45,46,17]. Finally, the predicted M/E sites that belonged to A-to-I, C-to-U and Other were manually examined.

All identified M/E sites were named by the names of the pre-miRNAs, positions of the sites, the nucleotides from the reference pre-miRNA sequences and the edited/mutated nucleotide at the sites.

### Comparing the M/E sites to reported SNPs

The identified M/E sites were compared to known SNPs in miRNAs organized in [47] (which was based on the dbSNP v137) and reported SNPs in the dbSNP (v151). Only sites that satisfied the following criteria were regarded as SNPs, (i) had the same genomic positions as the SNPs, (ii) had the same nucleotides as the alleles of the SNPs for both the original and changed nucleotides, and (iii) had editing levels of 100% in at least one of the 131 samples selected.

### Identifying conserved editing sites in miRNAs

The A-to-I and C-to-U editing sites were compared to their counterparts in *Macaca mulatta* [18] and *Mus musculus* [2,3]. The editing sites of the same editing types that locate on the same positions of mature miRNAs of at least two different species were considered as conserved editing sites.

### Identifying age-related miRNA editing sites

The Pearson correlation, as well as its *P* -value, between editing level (in %) of each of the 2456 editing sites in Table S1 and the age of death (y) was calculated with the corr function in MatLab (Mathworks, MA) for the 29 PD-PC and 36 PC samples, respectively. The editing sites with *P* -values smaller than 0.05 were regarded as age-related.

### Identifying M/E sites with significantly different editing levels in PD

The editing levels of 2456 M/E sites in the PD-PC and PC; PC-Am and AM samples were compared, respectively, with the Mann-Whitney *U* -test. The obtained *P* -values were corrected with the Benjamini-Hochberg correction method [41]. M/E sites with multiple test corrected *P* -value smaller than 0.05 were regarded as having significantly different editing levels in PD-PC or PC-Am samples.

### Identifying targets for original and edited miRNAs

M/E sites that satisfy the following criteria were chosen to identify the targets of original and edited miRNAs, (i) have significantly different editing levels in PD-PC samples compared to PC samples; and (ii) locate in the seed regions of mature miRNAs. The targets of original and edited miRNAs were predicted using the MiCPAR algorithm [20] with its default parameters. The targets with at least 1 PAR-CLIP read with T-to-C variation were kept in further analysis.

As listed in Table S1.3, seven PAR-CLIP sequencing profiles prepared from HEK293 cells stably expressing FLAG/HA-tagged AGO proteins (AGO1, AGO2, AGO3, and AGO4) [48] were downloaded from the NCBI SRA database using the series accession number SRP002487. Four PAR-CLIP sequencing profiles prepared from HEK293 cell lines stably expressing HIS/FLAG/HA-tagged AGO1 or AGO2 [49] were downloaded from the NCBI SRA database using the series accession number SRP018015. Raw reads in these profiles were filtered to make sure that the first 25 nucleotides of the qualified reads have sequencing scores of 30 or higher. The 3’ adapters were cut for qualified reads. The remaining reads in these 11 profiles were combined and used in the identification of miRNA targets with the MiCPAR algorithm [20]. The annotation of NCBI RefSeq genes in the GTF file, the mRNA sequences of NCBI RefSeq genes (refMrna.fa.gz, version hg38) and soft-masked genome sequences of human (version GRCh38) were downloaded from the UCSC Genome Browser [50] and used as inputs of the MiCPAR algorithm. The targets having at least 1 PAR-CLIP read with T-to-C variation were kept for further analysis.

### GO and Pathway analysis for the original and edited miRNAs

The GO term and KEGG pathway enrichment of the targets that were only targeted by the original or edited miRNAs were analyzed with KOBAS2, respectively [51]. The significantly enriched GO terms (with multiple test corrected *P* -values smaller than 0.05) were divided into Biological Process, Cellular Component and Molecular Function. Then, the enriched GO terms and KEGG pathways of the original and edited miRNAs were compared.

### Identifying meaningful targets for edited miRNAs

A previous study [21] identified 1095 deregulated genes with the Wald tests (in DESeq2 [52]) by analyzing RNA-seq data sets for 19 PD and 24 control samples of prefrontal cortex (Brodmann Area 9, BA9). This study also examined the protein abundance levels of BA9 samples of 12 PD patients and 12 normal people using MS3 proteomics and identified 283 proteins with significantly different levels (limma package, corrected *P* < 0.05) in PD [21]. Another study [53] generated 30 gene expression profiles of BA9 with microarray, with 15 profiles for PD and PD-PC respectively. The deregulated genes were identified with the limma package.

Because these RNA-seq and proteomics profiles were also from BA9 which is same as the regions of PD-PC and PC sRNA-seq profiles in this study, these deregulated genes (mRNAs) and proteins were used to identify meaningful targets for edited miRNAs. For edited miRNAs that have higher editing levels and normalized abundances in the PD-PC samples, we chose their targets, identified with the MiCPAR algorithm, which showed lower expression levels in PD-PC samples, and vise versa.

### Validating selected targets of edited hsa-miR-497-5p

The 3’ UTR segments of human OPA1/VAPB, mutated OPA1/VAPB, crab-eating monkey (*Macaca fascicularis*) OPA1/VAPB, mutated monkey OPA1/VAPB and mouse (*Mus musculus*) Opa1/Vapb containing the hsa-mir-497_25g complementary sites (*∼* 130 nt with 70 nt on both sides) were synthesized and cloned into the plasmid pmirGLO (Promega, Madison, WI, US) with XholI and SalI sites, and named as pGLO-OPA1/VAPB, pGLO-OPA1m/VAPBm, pGLO-mfaOPA1/mfaVAPB, pGLO-mfaOPA1m/mfaVAPBm, and pGLO-musOpa1/musVapb, respectively. The genomic regions of hsa-pre-mir-497 and hsa-pre-mir-497 25g were synthesized and cloned into the plasmid pCDNA 3.1(+) (Invirtrogen, Carlsbad, CA, USA) with HindIII and BamHI sites, and named as p497 and p497 25g, respectively. All the plasmids were validated by Sanger sequencing (ABI3730, Thermo Fisher, Waltham, MA, US).

Human 293T cells was cultured in DMEM high glucose medium with 10% FBS, 1% NEAA. 500 µL medium was inoculated to each well with 24 well plate. 2 µg p497 25g and 2 µg pGLO-OPA1 were added to one well as a group of co-transfection. Similarly, one pGLO plasmid and one pCDNA were co-transfected to 293T cells, respectively. Then added and mixed 2 µL of DNA-INVI DNA Transfection Reagent (Invigentech, Carlsbad, CA 92008, USA) in each well.

In the experiments, each group had three biological replicates. TransDetect double-luciferase Reporter Assay Kit (Beijing TransGen Biotech, Beijing, China) was used to detect luciferase activities for each of the biological replicates with three technical repeats. The mean value of the three technical repeats was used as the value of one biological replicate. The luciferase activities of different groups were then compared with two tailed *t*-tests.

### Conservation analysis of edited miR-497-5p complementary sites

The conservation scores (PhyloP scores for 30 mammals) of the A-to-I edited miR-497-5p complementary sites on OPA1 and VAPB were downloaded from UCSC Genome Browser [38]. The sequences of the A-to-I edited miR-497-5p complementary sites on OPA1 and VAPB in 30 mammals were downloaded from the UCSC Genome Browser and used to build phylogenetic trees with ClustalX (v2.1) [54], respectively. The obtained phylogenetic trees were visualized with TreeView (v1.6.6) [55].

## Data Availability

The data used in this study were available at the NCBI SRA and GEO databases at https://www.ncbi.nlm.nih.gov/sra and https://www.ncbi.nlm.nih.gov/geo/.

## Data Availability

The 131 sRNA-seq profiles, 7 cohorts of gene expression profiles, and 11 PAR-CLIP sequencing profiles were available at the NCBI GEO and SRA databases under the accession numbers listed in Table S1.

## Authors’ contributions

YZ conceived and designed the research. CL, SR, ZZ, XW, AS, NZ, JY, CP, and YZ analyzed the data and organized the results. CL, XW, SW and TL performed the luciferase experiments. YZ developed the computational pipelines for analyzing the data sets. YZ wrote the manuscript. All authors had read and approved this paper in the current form.

## Competing interests

The authors declare that they have no competing interests.

## Acknowledgements

The research was supported in part by a grant (No. 31760314) of National Natural Science Foundation of China (http://www.nsfc.gov.cn/) to YZ and a grant (No. 2018YFA0108502) of the Ministry of Science and Technology of China to YZ. The funders had no role in study design, data collection and analysis, decision to publish, or preparation of the manuscript.

## Additional information

Supplementary information is available for this paper at the website of the journal. Correspondence and requests for materials should be addressed to YZ.

## Notes

### Competing Interest Statement

The authors have declared no competing interest.

### Funding Statement

The research was supported in part by two grants (No. 31760314) of National Natural Science Foundation of China and a grant (No. 2018YFA0108502) of the Ministry of Science and Technology of China to YZ. The funders had no role in study design, data collection and analysis, decision to publish, or preparation of the manuscript.

### Summary of Updates

Add a new author, Shuai Wu, was added. Update name of author Xingwang Wu. Fig. 4 was updated. A new figure Fig. 9 was added in Extended Data. The related parts in Methods were also updated. Table S9 was updated. A new Supplementary Table, Table S19, was added.

