## Extended Data: Fig. 1 to Fig. 9 for "Characterizing relevant microRNA editing sites in Parkinson’s disease"

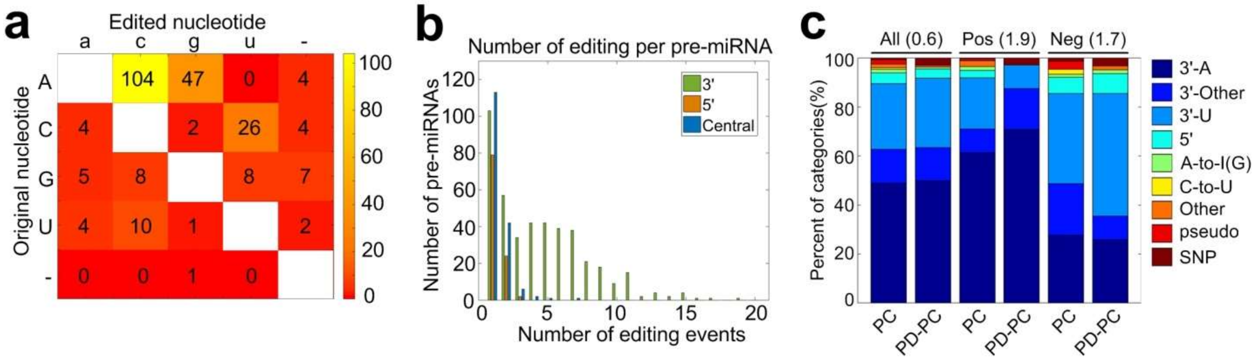


**Extended Data Fig. 1** The distribution of different types of editing sites in miRNAs. (**a**) The numbers of different types of editing events that do not happen at the 5' or 3' end of mature miRNAs. (**b**) The distribution of the numbers of pre-miRNAs with different numbers of 5'-, 3'- and Central editing sites, i.e. editing sites that do not happen at the ends of mature miRNAs. (**c**) The percentages of different types of miRNA editing sites whose editing levels are significantly correlated with the ages of individuals in PC and PD-PC samples. The values in the parenthesis are the Kullback-Leibler divergences between the distributions of PC and PD-PC. The source data are provided in Supplementary Table S10.

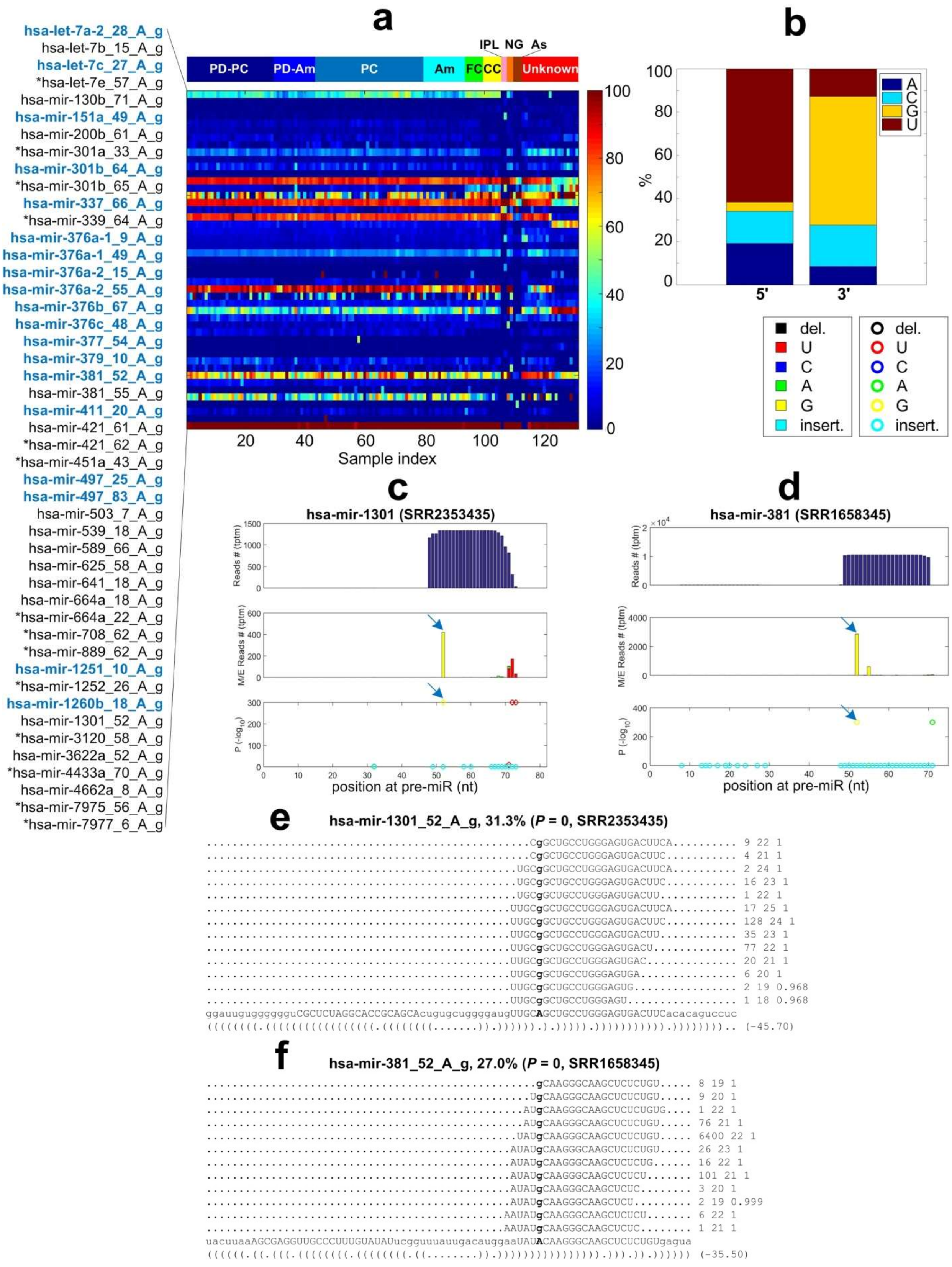
a
b
c
d
e
f
hsa-let-7a-2_28_A_g
hsa-let-7b_15_A_g
hsa-let-7c_27_A_g
*hsa-let-7e_57_A_g
hsa-mir-130b_71_A_g
hsa-mir-151a_49_A_g
hsa-mir-200b_61_A_g
*hsa-mir-301a_33_A_g
hsa-mir-301b_64_A_g
*hsa-mir-301b_65_A_g
hsa-mir-337_66_A_g
*hsa-mir-339_64_A_g
hsa-mir-376a-1_9_A_g
hsa-mir-376a-1_49_A_g
hsa-mir-376a-2_15_A_g
hsa-mir-376a-2_55_A_g
hsa-mir-376b_67_A_g
hsa-mir-376c_48_A_g
hsa-mir-377_54_A_g
hsa-mir-379_10_A_g
hsa-mir-381_52_A_g
hsa-mir-381_55_A_g
hsa-mir-411_20_A_g
hsa-mir-421_61_A_g
*hsa-mir-421_62_A_g
*hsa-mir-451a_43_A_g
hsa-mir-497_25_A_g
hsa-mir-497_83_A_g
hsa-mir-503_7_A_g
hsa-mir-539_18_A_g
hsa-mir-589_66_A_g
hsa-mir-625_58_A_g
hsa-mir-641_18_A_g
hsa-mir-664a_18_A_g
*hsa-mir-664a_22_A_g
*hsa-mir-708_62_A_g
*hsa-mir-889_62_A_g
hsa-mir-1251_10_A_g
*hsa-mir-1252_26_A_g
hsa-mir-1260b_18_A_g
hsa-mir-1301_52_A_g
*hsa-mir-3120_58_A_g
hsa-mir-3622a_52_A_g
*hsa-mir-4433a_70_A_g
hsa-mir-4662a_8_A_g
*hsa-mir-7975_56_A_g
*hsa-mir-7977_6_A_g
IPL NG As
PD-PC PD-Am PC Am FC CC Unknown
Sample index
A C G U
5' 3'
%
del. U C A G insert.
hsa-mir-1301 (SRR2353435)
hsa-mir-381 (SRR1658345)
Reads # (tptm)
M/E Reads # (tptm)
P (-log10)
position at pre-miR (nt)
hsa-mir-1301_52_A_g, 31.3% (P = 0, SRR2353435)
CgGCUGCCUGGGAGUGACUUCA 9 22 1
CgGCUGCCUGGGAGUGACUUC 4 21 1
UGCgGCUGCCUGGGAGUGACUUCA 2 24 1
UGCgGCUGCCUGGGAGUGACUUC 16 23 1
UGCgGCUGCCUGGGAGUGACUU 1 22 1
UUGCgGCUGCCUGGGAGUGACUUCA 17 25 1
UUGCgGCUGCCUGGGAGUGACUUC 128 24 1
UUGCgGCUGCCUGGGAGUGACUU 35 23 1
UUGCgGCUGCCUGGGAGUGACU 77 22 1
UUGCgGCUGCCUGGGAGUGAC 20 21 1
UUGCgGCUGCCUGGGAGUGA 6 20 1
UUGCgGCUGCCUGGGAGUG 2 19 0.968
UUGCgGCUGCCUGGGAGU 1 18 0.968
ggauuguggggguCGCUCUAGGCACCGCAGCAcugugcuggggaugUUGCAGCUGCCUGGGAGUGACUUCacacaguccuc
((((((((.(((((((((((((((((.((((((((.......)).)))))).).))))).))))))))))).)))))))).. (-45.70)
hsa-mir-381_52_A_g, 27.0% (P = 0, SRR1658345)
gCAAGGGCAAGCUCUCUGU 8 19 1
UgCAAGGGCAAGCUCUCUGU 9 20 1
AUgCAAGGGCAAGCUCUCUGUG 1 22 1
AUgCAAGGGCAAGCUCUCUGU 76 21 1
UAUgCAAGGGCAAGCUCUCUGU 6400 22 1
AUAUgCAAGGGCAAGCUCUCUGU 26 23 1
AUAUgCAAGGGCAAGCUCUCUG 16 22 1
AUAUgCAAGGGCAAGCUCUCU 101 21 1
AUAUgCAAGGGCAAGCUCUC 3 20 1
AUAUgCAAGGGCAAGCUCU 2 19 0.999
AAUAUgCAAGGGCAAGCUCUCU 6 22 1
AAUAUgCAAGGGCAAGCUCUC 1 21 1
uacuuaaAGCGAGGUUGCCCUUUGUAUAUucgguuuauugacauggaaUAUACAAGGGCAAGCUCUCUGUgagua
((((((.((.(((.((((((((.((((((((.((........)).)))))))))))))))).))).)).)))))) (-35.50)

**Extended Data Fig. 2** The details of 47 identified A-to-I editing sites in miRNAs. (**a**) The editing levels of the 47 A-to-I editing sites in the 131 selected brain data sets. The 14 sites marked with stars are newly identified sites. The 19 sites with blue names are conserved A-to-I editing sites in mammals. The 131 samples include 29 prefrontal cortex samples of postmortem PD patients (PD-PC), 14 amygdalae samples of postmortem PD patients (PD-Am), 36 prefrontal cortex samples of normal controls (PC), 14 amygdalae samples of normal controls (Am), 6 frontal cortex of normal controls (FC), 6 corpus callosum samples of normal controls (CC), 2 inferior parietal lobe samples of normal controls (IPL), 2 temporal neocortex gray matter samples of normal controls (NG), 3 astrocyte cell lines of normal controls (As), and 19 unknown brain regions of normal controls (Unknown). (**b**) The percentages of nucleotides beside the 47 A-to-I editing sites. (**c**) The MiRME map of hsa-mir-1301 in one of the PD-PC samples (SRR2353435). The upper panel shows the total numbers of reads that cover each nucleotide of the pre-miRNA. The central panel shows the numbers of M/E reads at each position of the pre-miRNA. And the lower panel givens multiple test corrected *P*-values (-log10 scaled) of the corresponding mutation/editing sites shown in the central panel. (**d**) The MiRME map of hsa-mir-381 in one of the unknown normal samples (SRR1658345). (**e**) The details of hsa-mir-1301_52_A_g in SRR2353435. (**f**) The details of hsa-mir-381_52_A_g in SRR1658345. In Part (e) and (f), the edited nucleotides are shown in bold face. The source data are provided in Supplementary Table S11.

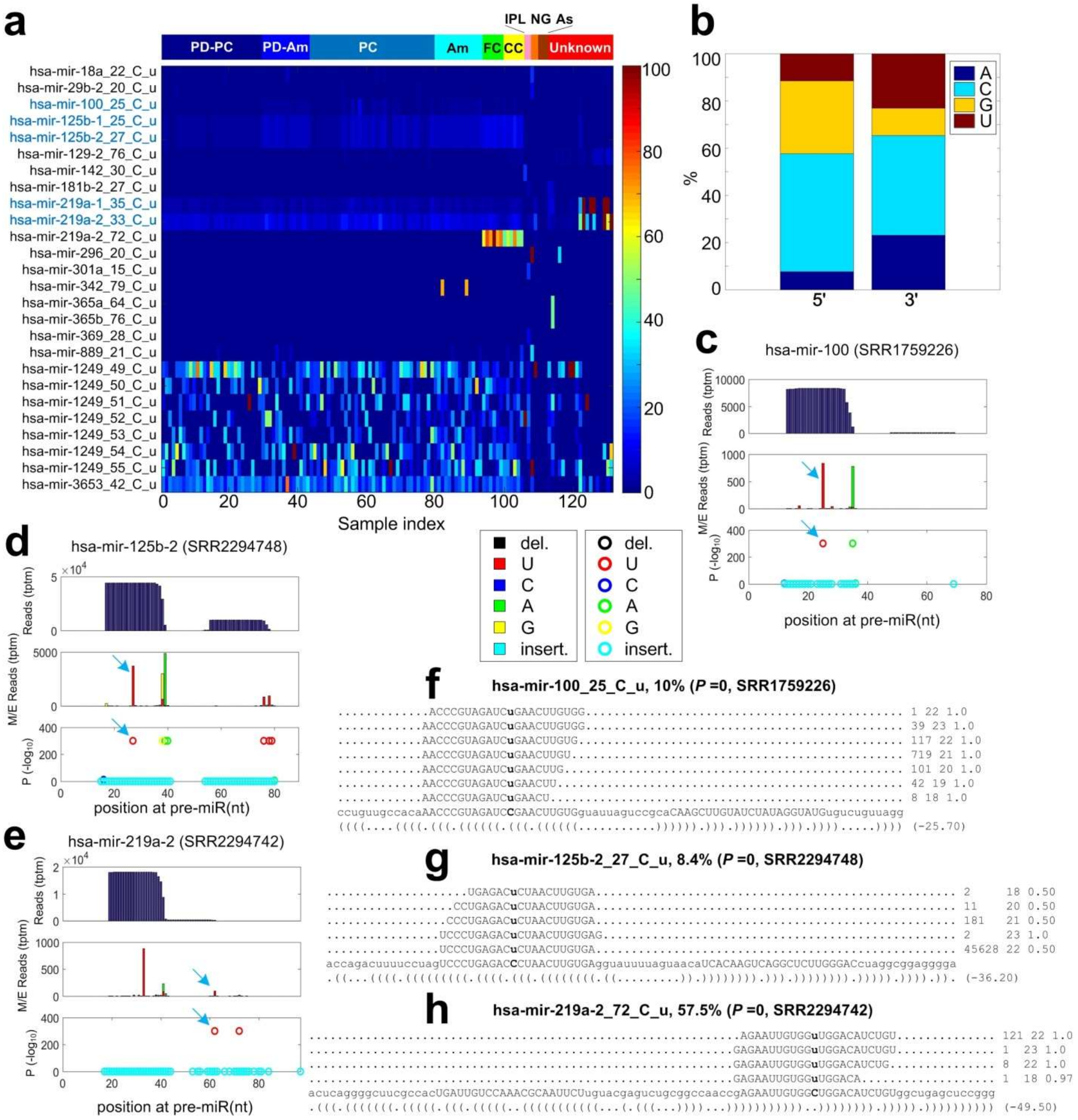


**Extended Data Fig. 3** The details of 26 identified C-to-U editing sites in miRNAs. (**a**) The editing levels of the 26 identified C-to-U editing sites in the 131 sRNA-seq profiles. The numbers of samples from different groups are the same as those in Extended Data Fig. 2a. The 5 sites with blue names are conserved C-to-U editing sites in primates. (**b**) The percentages of nucleotides beside these 26 C-to-U editing sites. (**c**) The MiRME map of hsa-mir-100 in one of the normal prefrontal cortex (PC) samples (SRR1759226). (c) The MiRME map of hsa-mir-125b-2 in one of the corpus callosum (CC) samples (SRR2294748). (**d**) The MiRME map of hsa-mir-219a-2 in one of corpus callosum (CC) samples (SRR2294742). (**e**) The details of hsa-mir-100_25_C_u in SRR1759226. (**f**) The details of hsa-mir-125b-2_27_C_u in SRR2294748. (**g**)

The details of hsa-mir-219a-2_72_C_u in SRR2294742. The source data are provided in Supplementary Table S12.

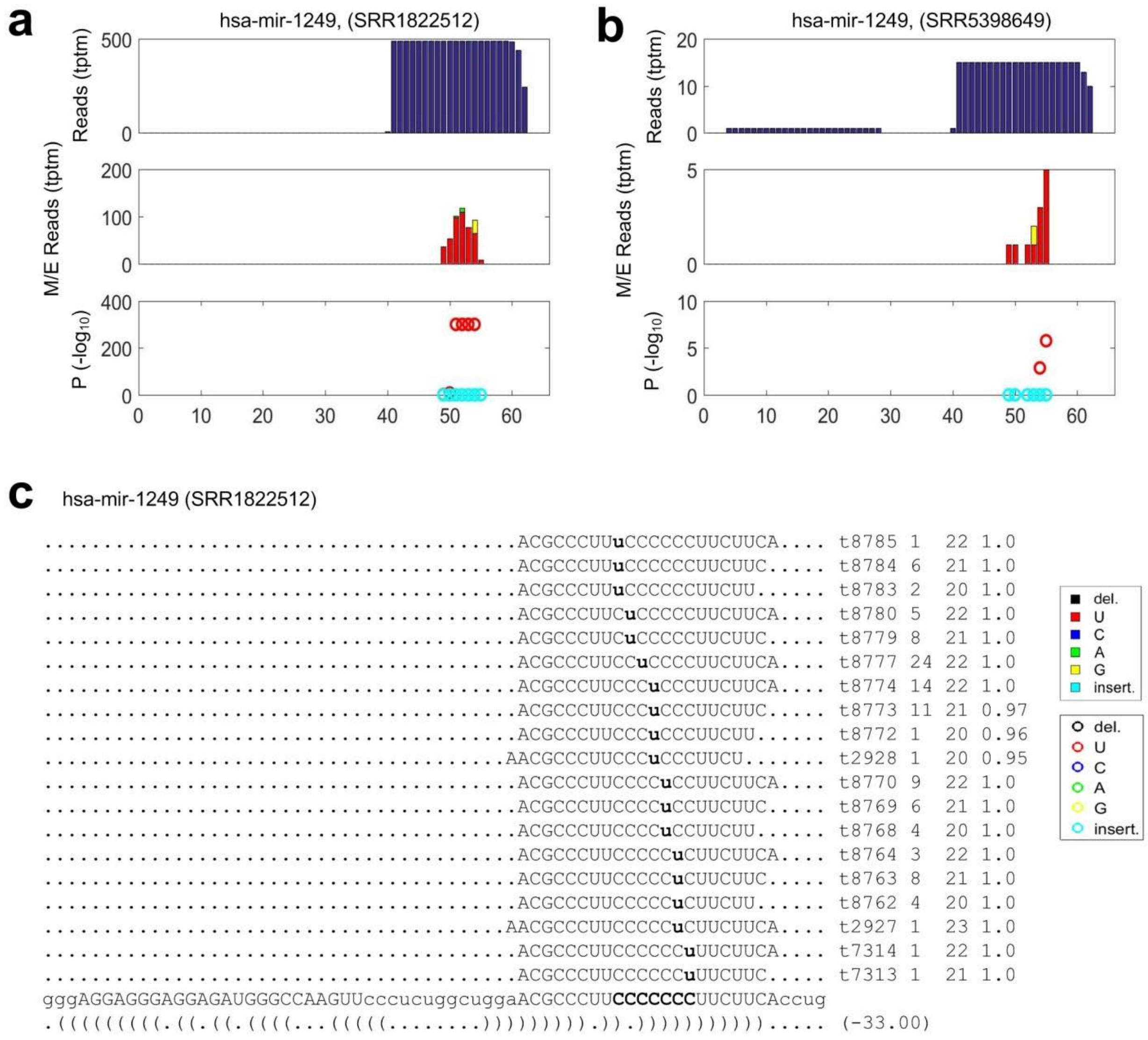


**Extended Data Fig. 4** Seven continuous C-to-U editing sites in hsa-mir-1249. (**a**) The MiRME map of hsa-mir-1249 in one of the inferior parietal lobe (IPL) samples (SRR1822512). (**b**) The MiRME map of hsa-mir-1249 in one of the amygdala (Am) samples (SRR5398649). (**c**) The details of the seven C-to-U editing sites in SRR1822512. The source data are provided in Supplementary Table S13.

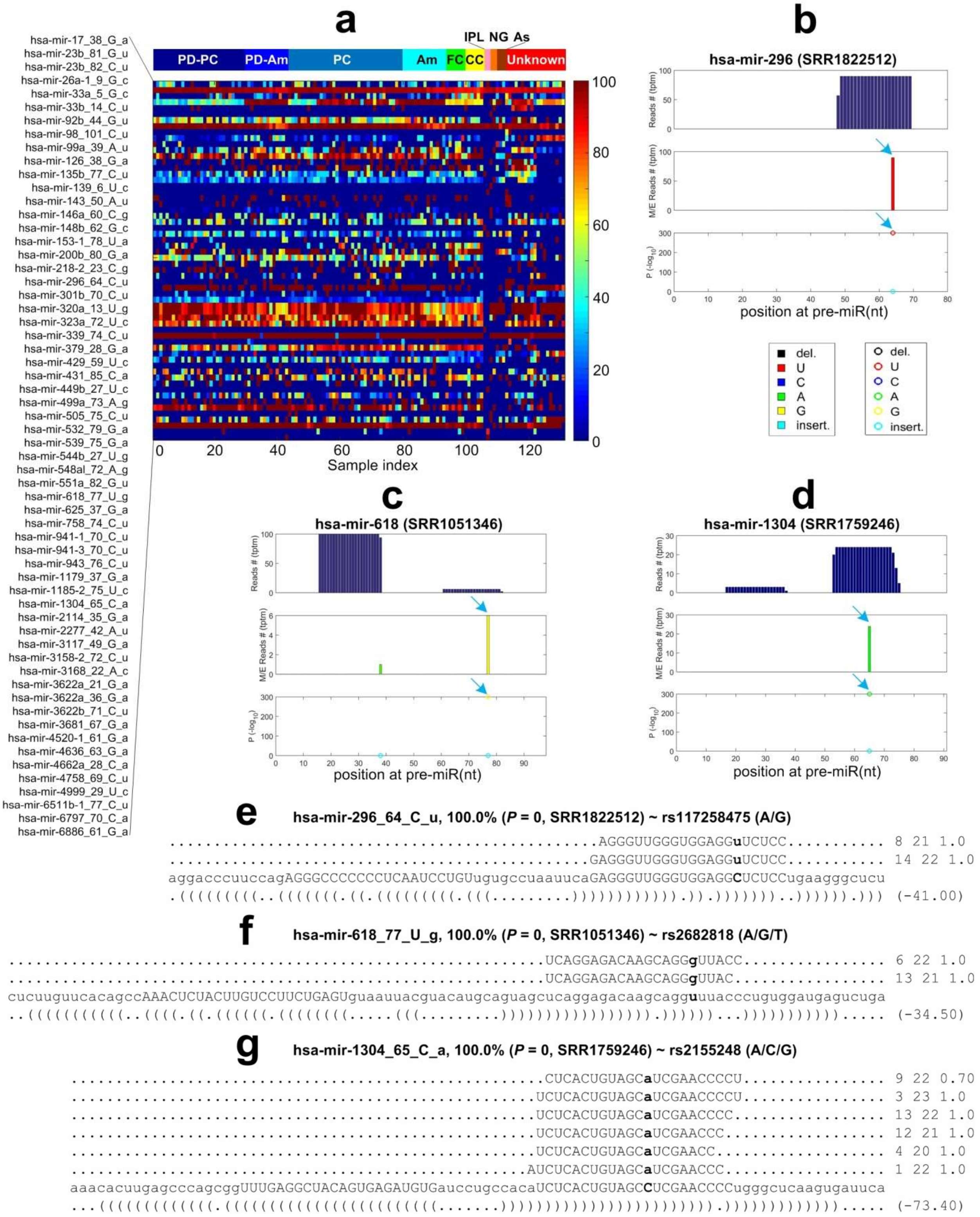


**Extended Data Fig. 5** The details of 60 identified SNPs in miRNAs. (**a**) The M/E levels of the 60 SNPs in the 131 sRNA-seq profiles. The numbers of samples from different groups are the same as those in Extended Data Fig. 2a. (**b**) The MiRME map of hsa-mir-296 in one of the

Inferior parietal lob (IPL) samples (SRR1822512). (**c**) The MiRME map of hsa-mir-618 in one of the unknown samples (SRR1051346). (**d**) The MiRME map of hsa-mir-1304 in one the normal prefrontal cortex samples (SRR1759246). (**e**) The details of hsa-mir-296_64_C_u, i.e., rs117258475, in SRR1822512. (**f**) The details of hsa-mir-618_77_U_g, i.e., rs2682818, in SRR1051346. (**g**) The details of hsa-mir-1304_65_C_a, i.e., rs2155248, in SRR1759246. The source data are provided in Supplementary Table S14.

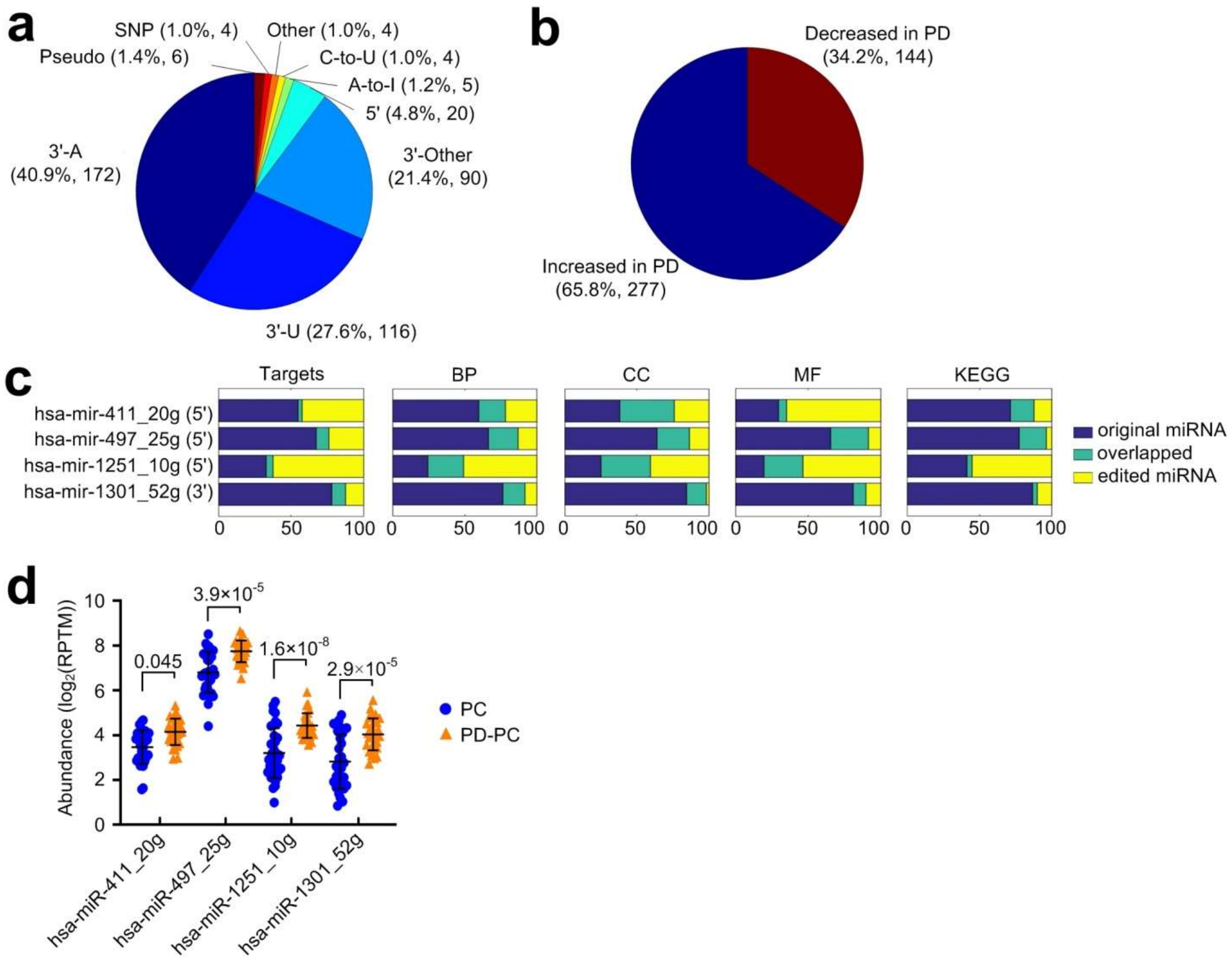


**Extended Data Fig. 6** The distribution of PD relevant editing sites. (**a**) The distribution of the different types of editing sites that have significantly different editing levels in PD-PC samples compared to the PC samples. (**b**) The distribution of sites in Part (a) whose editing levels are increased or decreased in PD-PC samples when compared to PC samples. (**c**) The comparisons of targets, GO terms, and KEGG pathways of original and four A-to-I edited miRNAs. The GO terms Biological Process (BP), Cellular Component (CC) and Molecular Function (MF) are shown separately. (**d**) The comparisons of normalized abundances of four A-to-I edited miRNAs in PC and PD-PC samples. The likelihood ratio tests (in edgeR) were used to compare the abundances of the edited miRNAs in different groups and to produce the *P*-values which were corrected with the Benjamini and Hochberg method. The multiple test corrected *P*-values were listed above the lines between PC and PD-PC samples. The source data are provided in Supplementary Table S15.

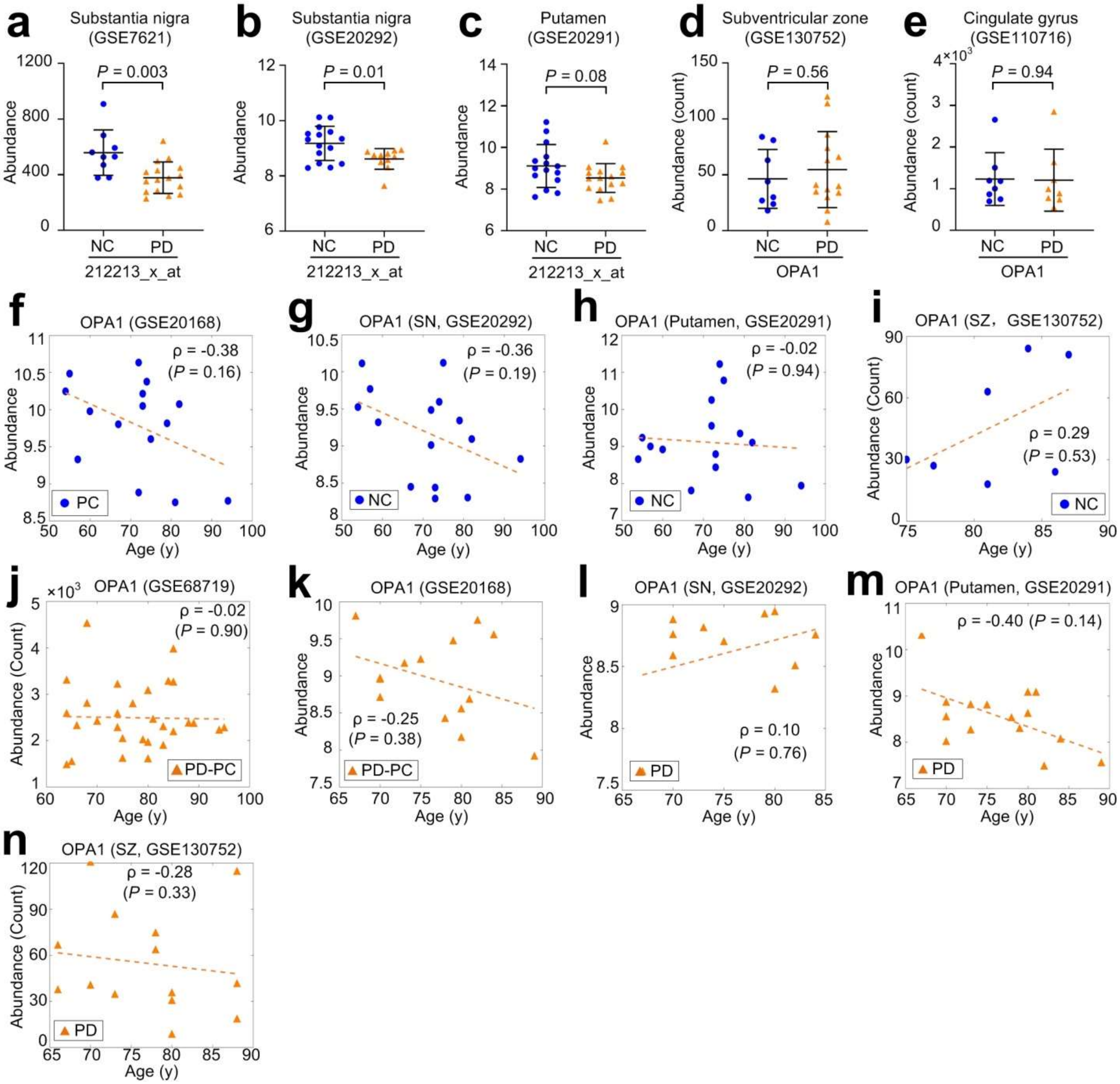


**Extended Data Fig. 7 The expression of OPA1 in different brain regions of PD patients and normal controls.** (**a**) The abundance of OPA1 in substantia nigra samples of Normal controls (NC) and PD patients (PD) (GSE7621). The *P*-value was calculated with the two-tailed *t*-test. (**b**) The abundance of OPA1 in substantia nigra samples of normal controls (NC) and PD patients (PD) (GSE20292). The *P*-value was calculated with the two-tailed *t*-test. (**c**) The abundance of OPA1 in putamen samples of normal controls (NC) and PD patients (PD) (GSE20291). The *P*-value was calculated with the two-tailed *t*-test. (**d**) The abundance of OPA1 in subventricular zone samples of normal controls (NC) and PD patients (PD) (GSE130752). The *P*-value was calculated with the two-tailed *t*-test. (**e**) The abundance of OPA1 in cingulate gyrus samples of normal controls (NC) and PD patients (PD) (GSE110716). The *P*-value was calculated with the two-tailed *t*-test. (**f**) The Spearman correlation (ρ) between the ages of PC samples and the abundances of OPA1 in GSE20168. (**g**) The Spearman correlation (ρ) between the ages of substantia nigra samples of normal controls (NC) and the abundances of OPA1 in GSE20292. (**h**) The Spearman correlation (ρ) between the ages of putamen samples of normal controls (NC) and the abundances of OPA1 in GSE20291. (**i**) The Spearman correlation between the ages of subventricular zone (SZ) samples of normal controls (NC) and the abundances of OPA1 in GSE130752. (**j**) The Spearman correlation (ρ) between the ages of PD-PC samples and the abundances of OPA1 in GSE68917. (**k**) The Spearman correlation (ρ) between the ages of PD-PC samples and the abundances of OPA1 in GSE20168. (**l**) The Spearman correlation between the ages of substantia nigra (SN) samples of PD patients (PD) and the abundances of OPA1 in GSE20292. (**m**) The Spearman correlation between the ages of the putamen samples of PD patients (PD) and the abundances of OPA1 in GSE20291. (**n**) The Spearman correlation between the ages of subventricular zone (SZ) samples of PD patients (PD) and the abundances of OPA1 in GSE130752. The source data are provided in Supplementary Table S16.

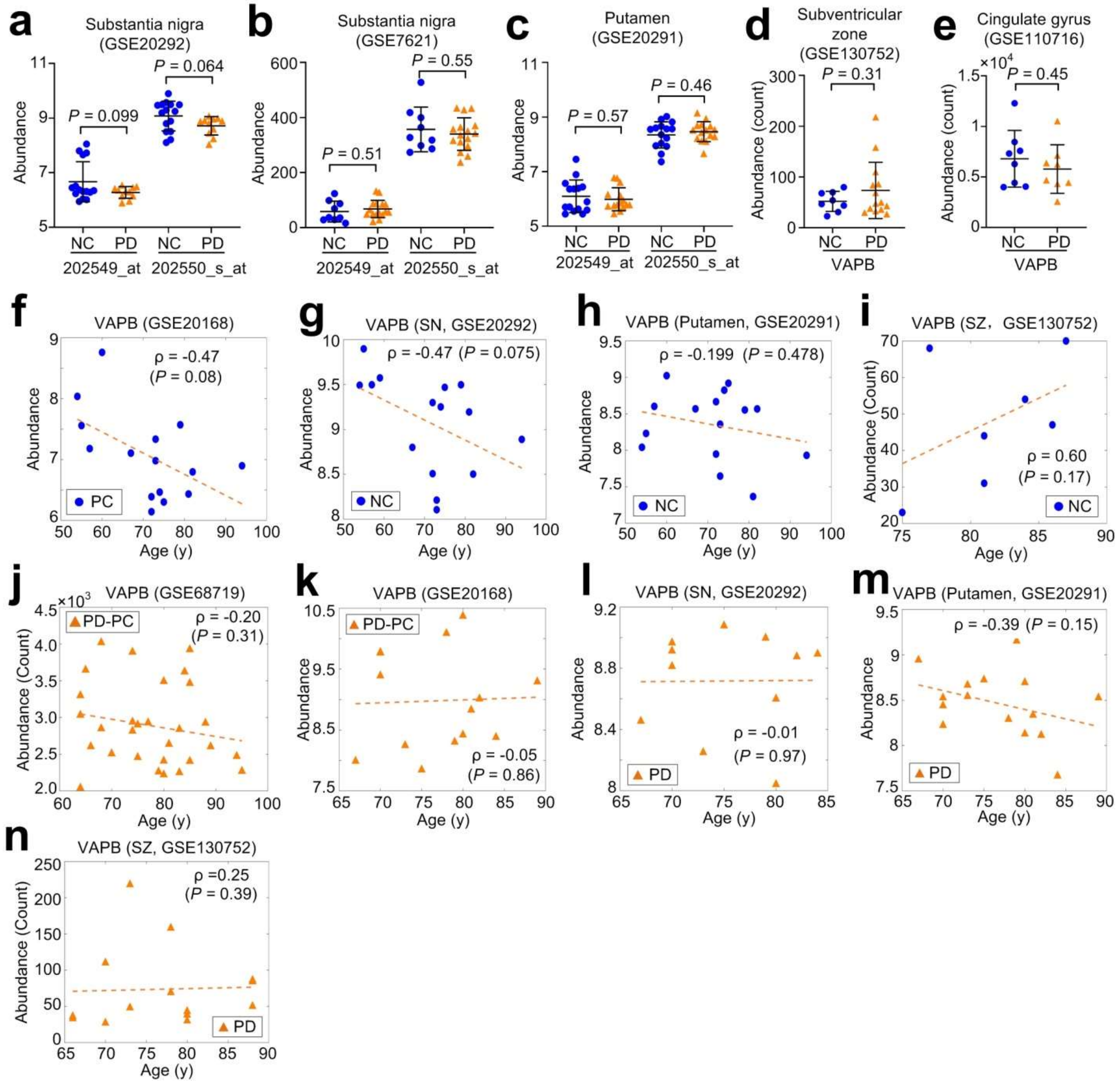


**Extended Data Fig. 8** The expression of VAPB in different brain regions of PD patients and normal controls. (**a**) The abundance of VAPB in substantia nigra samples of Normal controls (NC) and PD patients (PD) (GSE20292). The *P*-value was calculated with the two-tailed *t*-test. (**b**) The abundance of VAPB in substantia nigra samples of PD patients (PD) and normal controls (NC) (GSE7621). The *P*-value was calculated with the two-tailed *t*-test. (**c**) The abundance of VAPB in putamen samples of Normal controls (NC) and PD patients (PD) (GSE20291). The *P*-value was calculated with the two-tailed *t*-test. (**d**) The abundance of VAPB in subventricular zone samples of PD patients (PD) and normal controls (NC) (GSE130752). The *P*-value was calculated with the two-tailed *t*-test. (**e**) The abundance of VAPB in cingulate gyrus samples of PD patients (PD) and normal controls (NC) (GSE110716). The *P*-value was calculated with the two-tailed *t*-test. (**f**) The Spearman correlation (ρ) between the ages of PC samples and the abundances of VAPB in GSE20168. (**g**) The Spearman correlation (ρ) between the ages of substantia nigra samples of normal controls (NC) and the abundances of VAPB in GSE20292. (**h**) The Spearman correlation (ρ) between the ages of putamen samples of normal controls (NC) and the abundances of VAPB in GSE20291. (**i**) The Spearman correlation between the ages of subventricular zone (SZ) samples of normal controls (NC) and the abundances of VAPB in GSE130752. (**j**) The Spearman correlation (ρ) between the ages of PD-PC samples and the abundances of VAPB in GSE68917. (**k**) The Spearman correlation (ρ) between the ages of PD-PC samples and the abundances of VAPB in GSE20168. (**l**) The Spearman correlation between the ages of substantia nigra (SN) samples of PD patients (PD) and the abundances of VAPB in GSE20292. (**m**) The Spearman correlation between the ages of the putamen samples of PD patients (PD) and the abundances of VAPB in GSE20291. (**n**) The Spearman correlation between the ages of subventricular zone (SZ) samples of PD patients (PD) and the abundances of VAPB in GSE130752. The source data are provided in Supplementary Table S17.

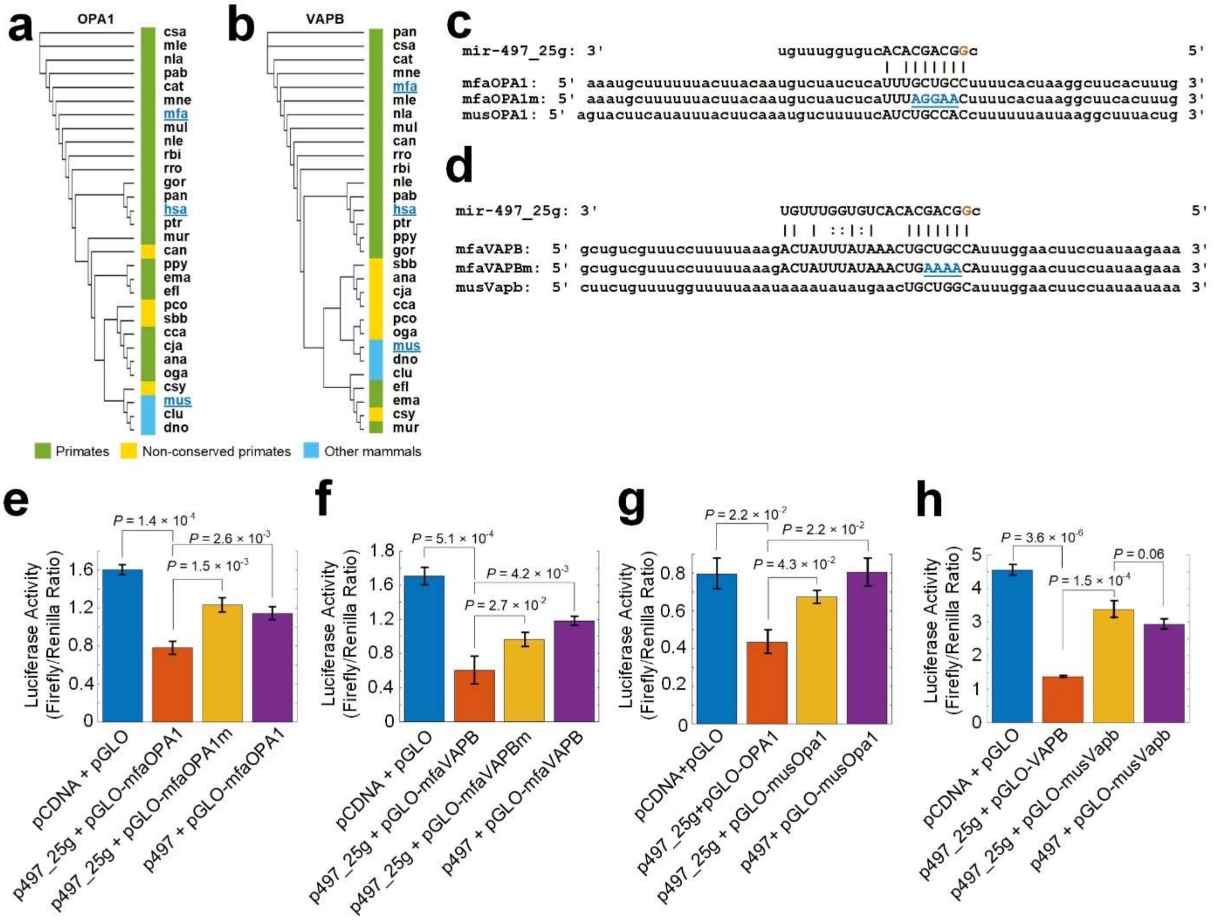

**Extended Data Fig. 9** Conservation and validation of OPA1 and VAPB as targets of A-to-I edited miR-497-5p. (**a**) Phylogenetic tree of A-to-I edited miR-497-5p complementary sites on OPA1 in 30 mammals. The abbreviations of the 30 species are listed in Supplementary Table S18. The three species with blue names, i.e., human (hsa, *Homo sapiens*), monkey (mfa, *Macaca fascicularis*) and mouse (mus, *Mus musculus*) were selected for luciferase experiments. (**b**) Phylogenetic tree of A-to-I edited miR-497-5p complementary sites on VAPB in 30 mammals. Legend are the same as those of Part (a). (**c**) The regions of monkey OPA1 (mfaOPA1), mutated monkey OPA1 (mfaOPA1m) and mouse OPA1 (musOPA1) with A-to-I edited miR-497-5p complementary sites. (**d**) The regions of monkey and mouse VAPB with A-to-I edited miR-497-5p complementary sites. (**e**) The luciferase activities when co-transfecting a pGLO plasmid of 3' UTRs of monkey OPA1 in Part (c) and a pCDNA plasmid containing original pre-hsa-mir-497 (p497) or pre-hsa-mir-497_25g (p497_25g), respectively. (**f**) The luciferase activities when co-transfecting a pGLO plasmid of 3' UTRs of monkey VAPB in Part (d) and a pCDNA plasmid containing original pre-hsa-mir-497 (p497) or pre-hsa-mir-497_25g (p497_25g), respectively. (**g**) The luciferase activities when co-transfecting a pGLO plasmid of 3' UTRs of mouse Opa1 in Part (c) and a pCDNA plasmid containing original pre-hsa-mir-497 (p497) or pre-hsa-mir-497_25g (p497_25g), respectively. (**h**) The luciferase activities when co-transfecting a pGLO plasmid of 3' UTRs of mouse Vapb in Part (c) and a pCDNA plasmid containing original pre-hsa-mir-497 (p497) or pre-hsa-mir-497_25g (p497_25g), respectively. In Part (e) to (h), the values shown are mean values ± SDs. *P*-values were based on two-tailed *t*-tests. The source data are available in Supplementary Table S18.
